# Estimating Real World Performance of a Predictive Model: A Case-Study in Predicting End-of-Life

**DOI:** 10.1101/19008821

**Authors:** Vincent J Major, Neil Jethani, Yindalon Aphinyanaphongs

**Author notes:** Correspondence to: Vincent J Major, 227 East 30th St, 6th Floor, New York, NY 10016.

## Abstract

**Objective:** The main criteria for choosing how models are built is the subsequent effect on future (estimated) model performance. In this work, we evaluate the effects of experimental design choices on both estimated and actual model performance.

**Materials and Methods:** Four years of hospital admissions are used to develop a 1 year end-of-life prediction model. Two common methods to select appropriate prediction timepoints (backwards-from-outcome and forwards-from-admission) are introduced and combined with two ways of separating cohorts for training and testing (internal and temporal). Two models are trained in identical conditions, and their performances are compared. Finally, operating thresholds are selected in each test set and applied in a final, ‘real-world’ cohort consisting of one year of admissions.

**Results:** Backwards-from-outcome cohort selection discards 75% of candidate admissions (n=23,579), whereas forwards-from-admission selection includes many more (n=92,148). Both selection methods produce similar global performances when applied to an internal test set. However, when applied to the temporally defined ‘real-world’ set, forwards-from-admission yields higher areas under the ROC and precision recall curves (88.3 and 56.5% vs. 83.2 and 41.6%).

**Discussion:** A backwards-from-outcome experiment effectively transforms the training data such that it no longer resembles real-world data. This results in optimistic estimates of test set performance, especially at high precision. In contrast, a forwards-from-admission experiment with a temporally separated test set consistently and conservatively estimates real-world performance.

**Conclusion:** Experimental design choices impose bias upon selected cohorts. A forwards-from-admission experiment, validated temporally, can conservatively estimate real-world performance.

## BACKGROUND AND SIGNIFICANCE

Performance estimation is a core evaluation for any prognostic or machine learning based model. Validation serves several related purposes: estimation of how the learned model performs when applied to new, unseen patients and selection of reasonable probability thresholds. Predictive models are typically validated internally with either subsampling methods [1] or explicit ‘hold out’ cohorts [2,3]. Internal validation is often the only validation used to estimate how the model may perform when implemented in practice.

Internal validation measures the generalization to unseen patients. A related quantity is model performance upon deployment into a similar but necessarily future population—sometimes called historic transportability [1]. One would expect these two quantities to be comparable, especially when the training distribution resembles that seen upon implementation. However, this is not always the case.

The tacit goal of any experimental design is to build a training set that closely resembles deployment. This constraint ensures that a model will work as intended and minimize introduced bias. In this paper, we revisit several methods to design experiments with retrospective patient data and to split data into model development sets. Within a setting of end-of-life prediction, we describe how experimental choices simplify and sanitize model development data and ultimately affect validation and estimated real-world performance.

### End-of-Life Use-Case

End-of-life interventions—those that are more palliative than curative—are often initiated too late in a patient’s life. Guidelines suggest palliative care is appropriate for anyone living with a chronic or serious illness that will eventually cause their death [4]. However, palliative care is often only initiated in the last weeks or months of life [5]. One reason for this may be that physicians are poor at estimating prognosis, typically being optimistic [6–8], influencing care decisions [9]. Practical systems that identify patients that may benefit from palliative care, or other end-of-life interventions, are needed [10,11].

Several recent works have described various machine learning methods to predict mortality as a proxy task for palliative care needs [2,12,13]. However, the experimental designs employed each have limitations. Namely, the validation cohorts no longer resemble the real-world cohort expected upon deployment.

## OBJECTIVE

The goal of this work is to compare several experimental design choices in terms of: 1) their effect on sanitizing model development data, and 2) the resulting predictive performance of a machine learning model. In particular, performance within two test cohorts will be compared and contrasted against that of a single, uniform ‘real-world’ cohort that, unlike the development data, is not affected by any experimental choices.

## MATERIALS AND METHODS

### Data

#### Prediction Task

Mortality prediction is a common task, but challenges in modeling, experimental design, and implementation have limited translation into practice. Mortality modeling works often introduce mortality risk as a proxy for patient appropriateness for palliative care [2,12]. In these proposed workflows, prediction timing is crucial. Inpatient admissions are promising as specialized clinicians are available to initiate meaningful interventions. More specifically, prediction at, or near, admission can aid the care team to approach the entirety of a patient’s admission cognizant of their risk.

Machine learning based approaches to mortality prediction often simplify ‘time-to-event’ analyses into binary outcomes of death within a certain time, commonly 1 year. As an objective for this work, we build on prior work and choose 1 year mortality as a target prediction task [2].

#### Patient Cohort

Data collected in routine clinical care of one urban, tertiary hospital is considered. Data is extracted from the clinical data warehouse where data is available from 2011. Admissions starting January 1, 2013 were considered for this work to allow prior patient data to accrue. At the time of data retrieval (March 2019) we consider admissions up to December 31, 2017, yielding five consecutive years of admissions as described in Figure 1A. However, the entire year of 2017 is held out of model development and will be used to simulate deployment.

**Figure 1.**
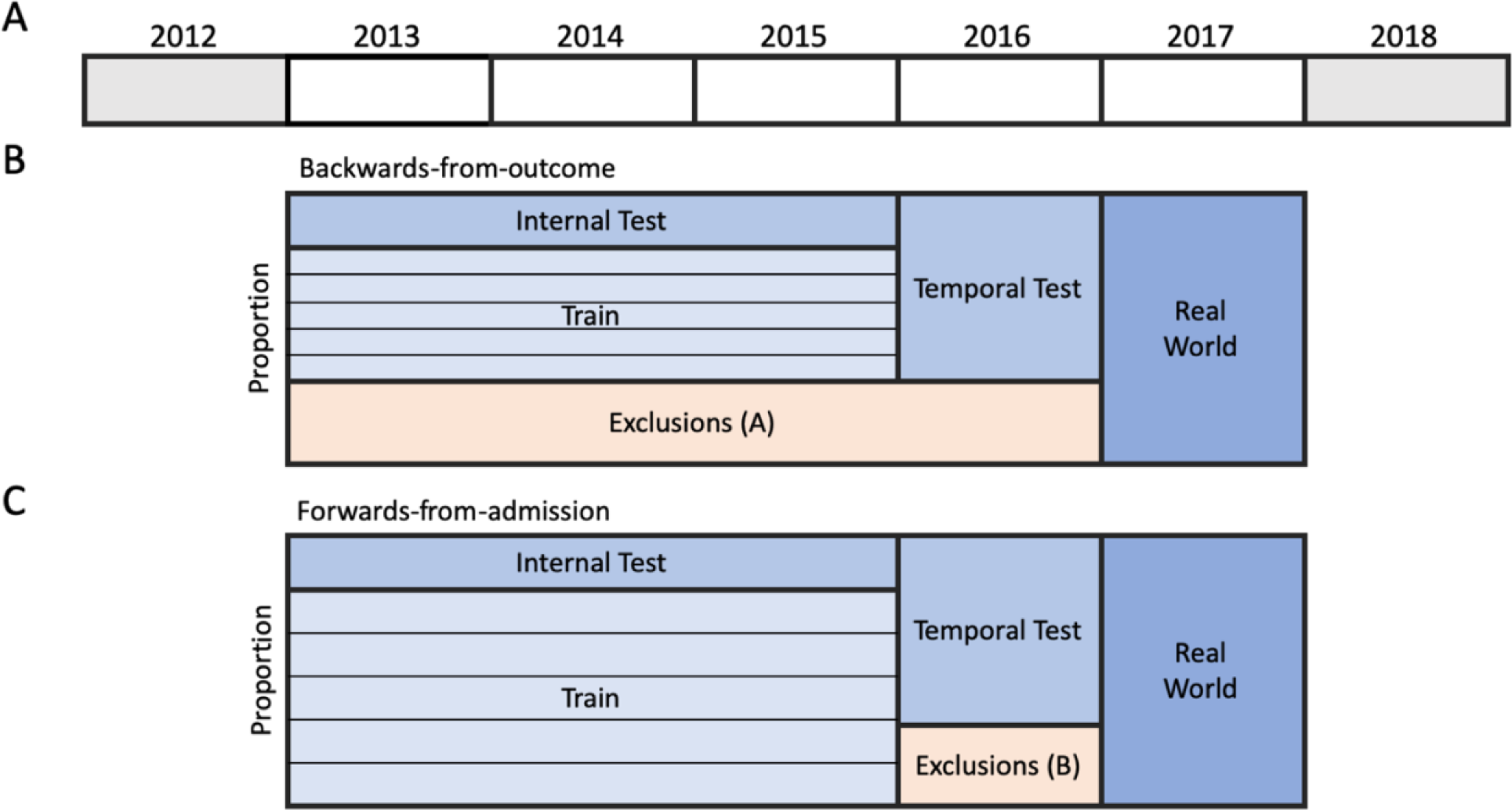
Schematic representation of available data and selected patient cohorts. A. 7 years of available data leaves 5 calendar years of admissions. B. Backwards-from-outcome cohort selection that only includes one admission per patient. C. Forwards-from-admission cohort selection that includes readmissions within each set but excludes readmissions into another set. The thin horizontal lines inside the training cohort depict cross-validation folds employed for parameter tuning.

#### Outcome Data

Mortality data is innately noisy as both the mechanism of death and effect of any life-preserving interventions vary between patients. Moreover, data collection is complicated by the inability to observe deaths beyond our hospital system. Three sources of data are combined to form a composite measure: 1) internal medical center death data, 2) purchased death data (derived from the Social Security Administration’s Master Death File) and 3) hospice discharges. None are perfect data capture mechanisms but together provide an ‘end-of-life’ label where the majority of patient outcomes are affirmed by at least two sources. We report a 8.5% prevalence of 1 year end-of-life in our cohort, comparable to other large prognostic cohorts [14].

#### Data Categories

The same broad data categories as prior works [2,9] are considered as predictors: demographics, encounters, diagnoses, medications, procedures, and laboratory results. Each of these categories, except demographics and encounters, have structured ontology-based vocabularies (ICD-10 diagnoses, RxNorm medications, CPT procedures, and LOINC laboratory tests). Data of this type can be problematic as there are a very large number of highly specific codes where only a few codes are attributed to each encounter. From a modeling perspective, code based data produces many sparse features which limits the number of feasible methods.

### Experimental Factors to Compare

Design of a retrospective experiment requires numerous choices. In this study, we introduce two particularly important decisions and compare two common approaches within each. Results from each of the four combinations will be presented similarly to enable direct comparisons.

#### Cohort Selection: by Outcome or Admission?

Due to the complexity of hospital admission data machine learning practitioners often subset the complete dataset to form a model development cohort using inclusion and exclusion criteria. The result is simplified and sanitized data that mitigate known challenges. Here, two contrasting experimental designs are presented: one that selects (instances) backwards from outcome and another that selects forwards from admission.

*Backwards-from-outcome cohort selection* refers to a strategy that starts from a known outcome and works backwards to identify an appropriate prediction instant for each patient, in effect, a retrospective case-control study. This design is also known as right-censoring. This method sanitizes the raw clinical data by excluding two particular challenges, uncertain labels, due to patients who are lost to follow-up, and 2) multiple admissions per patient, where the 1 year end-of-life outcome may flip from negative to positive.

Avati et al. [2] employ a backwards-from-outcome design to select one prediction instant per patient by working backwards from either 1) their known death, or 2) their last encounter. However, the authors enforce other criteria, namely, requiring one year of patient history. With such a design, Avati et al. select a cohort of 221,284 patients from a population of approximately two million [2]. These selection criteria are reproduced in this work.

*Forwards-from-admission cohort selection* refers to a design oriented forwards from each admission, analogous to recruitment of a prospective cohort study, sometimes referred to as left-censoring. The result is a cohort followed from their admission date without foresight of their outcome. Naturally, some patients are lost to follow-up within the 1 year observation period, or are subsequently readmitted. One advantage of forwards-from-admission selection is the more intuitive inclusion of readmissions, enabling a patient’s risk to evolve over time.

#### Test Set Selection: Internal Sample or Temporal Separation?

To evaluate how a model performs on unseen patients, a test set must be selected from the complete population before model development. How the test set is separated from the population determines how explicitly test set performance estimates both generalizability and historic transportability. Here, two common methods of selecting a test set are compared:

##### Internal Test Set

An internal test set includes patients randomly sampled from the same population and time period as the training data. In doing so, this test set measures generalizability. This design assumes sampling will ensure the testing and training distributions are similar, which may not hold in practice. Several recent mortality prediction works employ an internal test set in their work [2,12,13].

##### Temporal Test Set

A temporal test set includes patients from the same population but selects patients that are separated temporally from the training data. In doing so, this type of test set measures the historic transportability of the model into the near future, likely more challenging than generalizing. This test set is more closely aligned with the deployment process of training on prior data and deploying on new patients at some future time.

#### Combining Cohort Selection and Test Sets

Both backwards-from-outcome and forwards-from-admission experiments enable internal and temporal test sets. As depicted in Figure 1, a patient cohort is selected for both experiments and from which internal and temporal test sets are selected, leaving a training cohort.

Each individual patient should only exist in one cohort to mitigate data ‘leakage’ from training to testing. Backwards-from-outcome selection is simple as it yields one prediction per patient, whereas forwards-from-admission selection is more complex. In this work, only new patients in 2016 are recruited to the temporal test set. Excluded admissions are depicted in Figure 1B and 1C. Backwards-from-outcome selection removes all admissions except one that is closest to 1 year after working back from death or last censor. In contrast, selection forwards-from-admission removes a smaller set of readmissions from the temporal set to mitigate data leakage.

### Experimental Factors Consistent Across Designs

#### Feature Construction and Modeling

We fix the feature construction and modeling for each experimental design, by extending that of Avati et al. [2]. Specifically, the codes from each coded data type (diagnoses, medications, procedures, and lab results) are used in conjunction with the demographic and encounter information. A data observation window consists of the 365 days prior to admission plus data collected on hospital day 1. Features were constructed by splitting this time period into four ‘observation slices’ [2] where the codes within each are aggregated into:

- Count of unique codes across days,
- Count of total codes across days, and
- Mean, variance, minimum, maximum, and range of the number of codes assigned in a day.

Of note, LOINC lab result codes were included after separation into normal and abnormal groups which were aggregated independently. Infrequent features were subsequently pruned by removing codes that occurred in less than 100 records in each training set [2].

A random forest algorithm was employed, in particular the FEST implementation [15], for its benefits of robustness, high performance, and fast training especially with high dimensional, sparse data. Random forests tend to be reasonably insensitive to their parameters, nonetheless grid-search parameter tuning was conducted inside 5-fold cross-validation (sampled by patient to prevent data leakage) to select reasonable parameters before retraining one model on the entire training data. The final model is then applied to both the internal and temporal test sets.

#### Thresholding Criteria and Simulating Deployment

Typically, only metrics of overall model performance are reported—most commonly area under the receiver operating characteristic (AUROC) and, increasingly, area under the precision recall curve (AUPRC). However, in many applications an operating threshold is selected within a test set by imposing a criterion on one particular measure. Since our outcome of interest, end-of-life, is rare but very important, we are most concerned with systems operating at a clinically acceptable, typically high, precision (positive predictive value) [2] while maximizing recall (sensitivity). Together, these metrics can assist thresholding and inform how a model could be incorporated into an existing clinical workflow to recommend an intervention.

Deployment typically requires one operating threshold to separate patients at high-risk of dying from those at low-risk. We simulate this process by selecting a variety of potential operating thresholds by requiring precision to exceed some criterion (namely, the median selected threshold under bootstrapped subsampling conditions). We select 8 precision values spanning the range of realistic choices: 20–90%. Each threshold is ‘deployed’ by predicting in the real-world cohort and identifying patients who exceed the threshold. Notably, the real-world set is identical across experiments as it is not constrained to the same criteria as the development sets and is intended to represent the real-world where:

1. Patients are readmitted,
2. Patients have varying degrees of clinical history available,
3. Patients die within days of admission with no recent hospitalizations, and
4. Some patients are lost to follow-up and thus do not have a reliable outcome.

The first three factors are incorporated as sources of realistic noise but the last issue is difficult to overcome without introducing bias. For this cohort, patients lost to follow-up within 1 year are omitted.

As each threshold is applied, the performance criterion selected for within the test set produces a corresponding real-world performance. Direct comparison of model performance across test sets is unfair, as the groups are dissimilar, but the degree of ‘migration’ from test to real-world performance is reported and compared. Moreover, since the real-world cohort is consistent across experiments, the performance can be compared across thresholds, cohort selection method, and employed test sets.

## RESULTS AND DISCUSSION

### Patient Cohort

The study period spanned five calendar years, 2013–2017, and included all adult inpatient admissions to one hospital. In this period 128,328 admissions were recorded covering 87,293 unique patients. In this cohort the median [IQR] age at admission was 56.0 [35.4, 72.4] years, where 60.6% are female, 9.8% identify as Hispanic, 67.1% as white, 10.0% as African American, 8.1% as Asian, and 14.8% as other or unknown race. Of these admissions, 16,004 have a known death outcome (6,501 unique patients), with a median [IQR] time from admission to death of 138 [22, 493] days, where 10,932 admissions lead to end-of-life within 365 days, a prevalence of 8.5%.

The real-world cohort is separated early from this larger cohort. Defined as any admission in 2017 (n=29,382) but further restricted to patients known to have died or censored beyond 365 days results in a real-world cohort of 17,868 admissions. The model development period of 2013–2016 consists of 98,946 candidate admissions for cohort selection.

### Cohort Selection

#### Backwards-from-outcome Selection

The backwards-from-outcome experiment is very restrictive and discards the large majority of observed admissions yielding a model development cohort of n=23,579 that is separated into training, internal and temporal test sets as described in Table 1. The exclusions introduced by this design are apparent when plotted alongside the real-world set in Figure 2 (left). This approximate 75% reduction in cohort size is comparable to similar works, namely Avati et al.[2] that reported an approximate 90% reduction (221,284 patients from approximately 2 million).

**Table 1.** Model training and testing cohorts, stratified by outcome, for backwards-from-outcome and forwards-from-admission designs.

| Design | Outcome | Train | Internal Test | Temporal Test | Subtotal | Real-world |
| --- | --- | --- | --- | --- | --- | --- |
| Backwards-from-outcome | Survival | 11540 | 2906 | 7861 | 22307 | 15428 |
|  | Death | 732 (5.96%) | 192 (6.20%) | 348 (4.24%) | 1272 (5.39%) | 2440 (13.7%) |
| Forwards-from-admission | Survival | 52834 | 13389 | 18474 | 84697 | 15428 |
|  | Death | 4913 (8.51%) | 1157 (7.95%) | 1381 (6.96%) | 7451 (8.09%) | 2440 (13.7%) |

**Figure 2.**
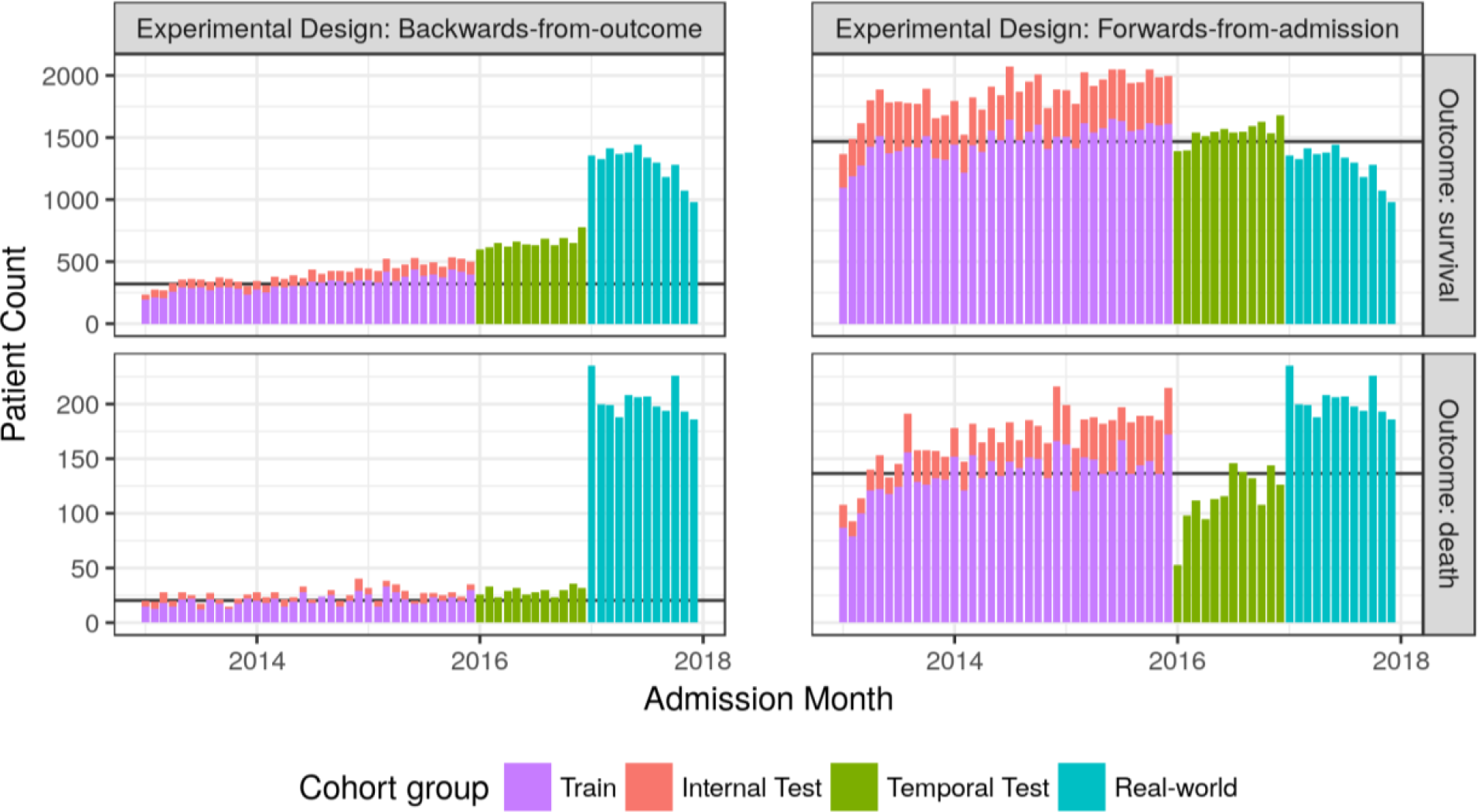
Cohort distributions of groups by calendar month. For reference, the monthly mean of the training set is represented with a horizontal black line.

One primary limitation of backwards-from-outcome selection is the introduction of temporal bias. Deaths occur reasonably uniformly across time, and thus selection should select similar number of deaths each month. However, when applied to the last encounter of the survival group, recent times are more likely to be selected as patients tend to continue to receive care within the same system until death, recovery or relocation. In the four years of model development data described in Figure 2, a noticeable increase in admissions per month is observed in the survival group (top-left) but not the death group (bottom-left). This introduced discrepancy between outcome groups will only grow as the time period extends, challenging the use of such a design in cohorts that span longer time periods.

#### Forwards-from-admission Selection

The less strict selection criteria of the forwards-from-admission design results in the inclusion of many thousands more admissions, evident in both Table 1 and Figure 2. The only admissions removed are those excluded upon transition to temporal test, where a noticeable drop is evident in Figure 2 starting January 2016, particularly in the death group (bottom-right). The resultant cohort of 92,148 admissions is almost four times larger than the backwards-from-outcome cohort with a larger mortality prevalence, closer to that of the population.

### Test Set Performance: Internal vs. Temporal

#### Backwards-from-outcome Selection

Parameter optimization in 5-fold cross-validation within the backwards-from-outcome training set selected the optimal random forest model with 500 trees, depth of 100, and negative sample weight of 0.2. The mean AUROC and AUPRC within cross-validation is 89.9% and 41.0% respectively. When retrained and applied to test sets, model performance shifts unpredictably as reported in Table 2, highlighting the differences between the two test sets.

**Table 2.** Model development and testing cohort performances in terms of AUROC and AUPRC for backwards-from-outcome and forwards-from-admission designs.

| Design | Evaluation Measure | Train (mean cross-validation) | Internal Test | Temporal Test | Real-world |
| --- | --- | --- | --- | --- | --- |
| Backwards-from-outcome | AUROC (%) | 89.9 | 89.5 | 85.4 | **83.2** |
|  | AUPRC (%) | 41.0 | 45.9 | 29.3 | **41.6** |
| Forwards-from-admission | AUROC (%) | 90.4 | 90.5 | 90.3 | **88.3** |
|  | AUPRC (%) | 48.7 | 45.0 | 46.9 | **56.5** |

Precision recall curves are plotted in Figure 3 for internal (left; red) and temporal (right; green) test sets alongside that of the real-world cohort (blue). Performance appears better when evaluating the model on the internal test set as compared to the temporal test set. Each operating threshold migrates from the estimated test set performance into a real-world performance, depicted by connected colored dots. Importantly, these performance migrations are not small vertical or horizontal movements, instead, they describe drastic performance shifts in both precision and recall. Interestingly, both internal and temporal test set precision recall curves intersect the real-world curve such that the direction of the performance shift depends on the desired precision criteria. These large shifts suggest that **neither test set provides an accurate estimation of real-world performance when using a backwards-from-outcome design**, likely due to distributional differences introduced by right-censoring.

**Figure 3.**
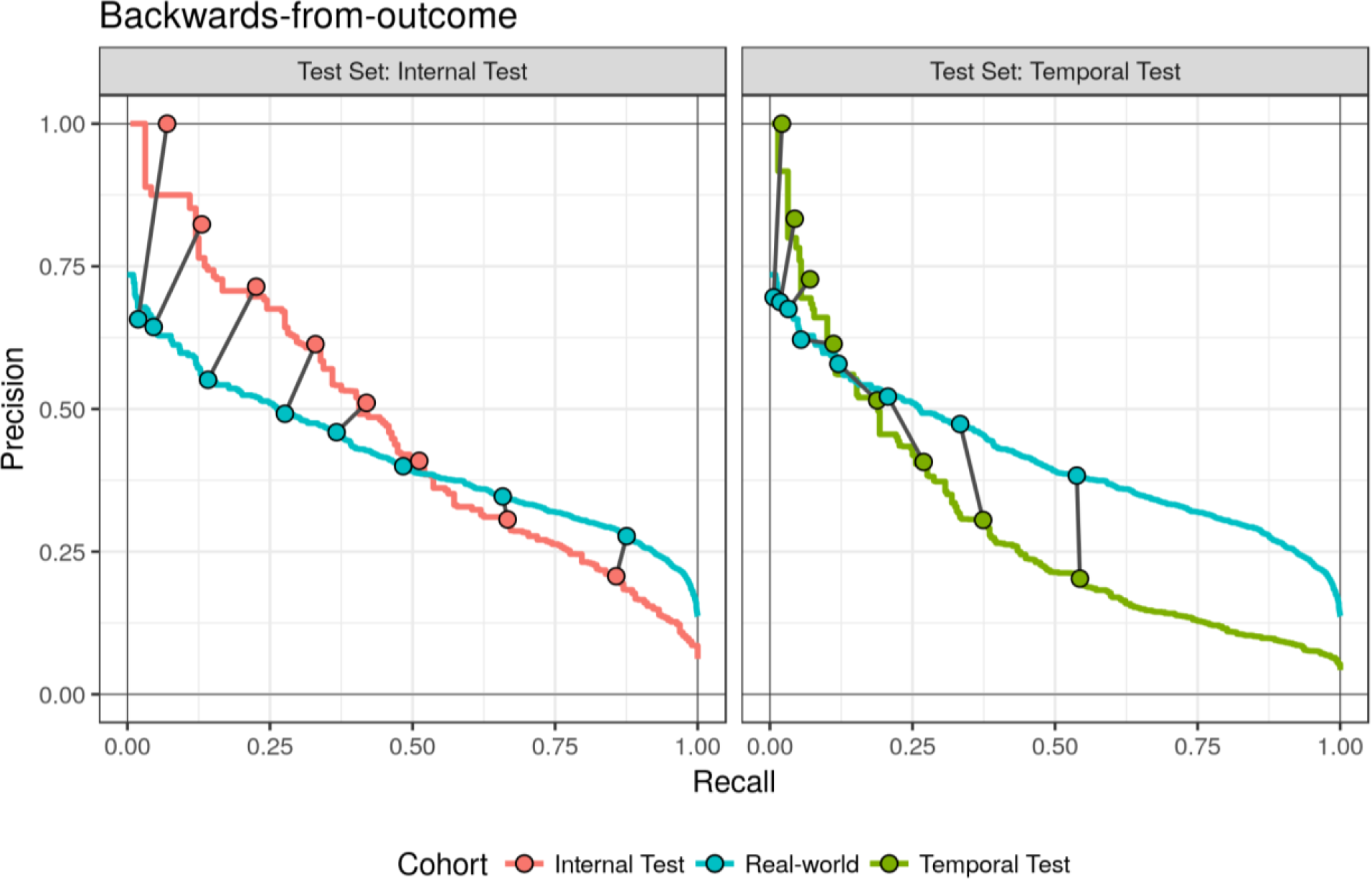
Precision and recall curves with highlighted operating points describing estimated real-world performance by the **backwards-from-outcome** experimental design in comparison to the internal and temporal test sets.

#### Forwards-from-admission Selection

Parameter optimization in 5-fold cross-validation within the forwards-from-admission training set selected the optimal random forest model with 1000 trees, depth of 200, and negative sample weight of 0.2, similar to the backwards-from-outcome design. The mean AUROC and AUPRC within cross-validation is 90.4% and 48.7% respectively and only marginal reductions in performance are reported when applied to internal and temporal test sets, described in Table 2, suggesting the cohorts are similar.

Both the internal and temporal test set precision recall curves of Figure 4 describe a drastic drop in performance in the very low recall region—known as the early retrieval problem [16] that is especially common in tasks with imperfect labels. Despite this drop in precision, thresholds selected in both test sets typically result in improved performances when applied to the real-world set. The performance difference is relatively consistent between each test set and the real-world set, but **employing a forwards-from-admission design with a temporal test set provides consistently conservative estimation of deployment—**a safe scenario for deployment.

**Figure 4.**
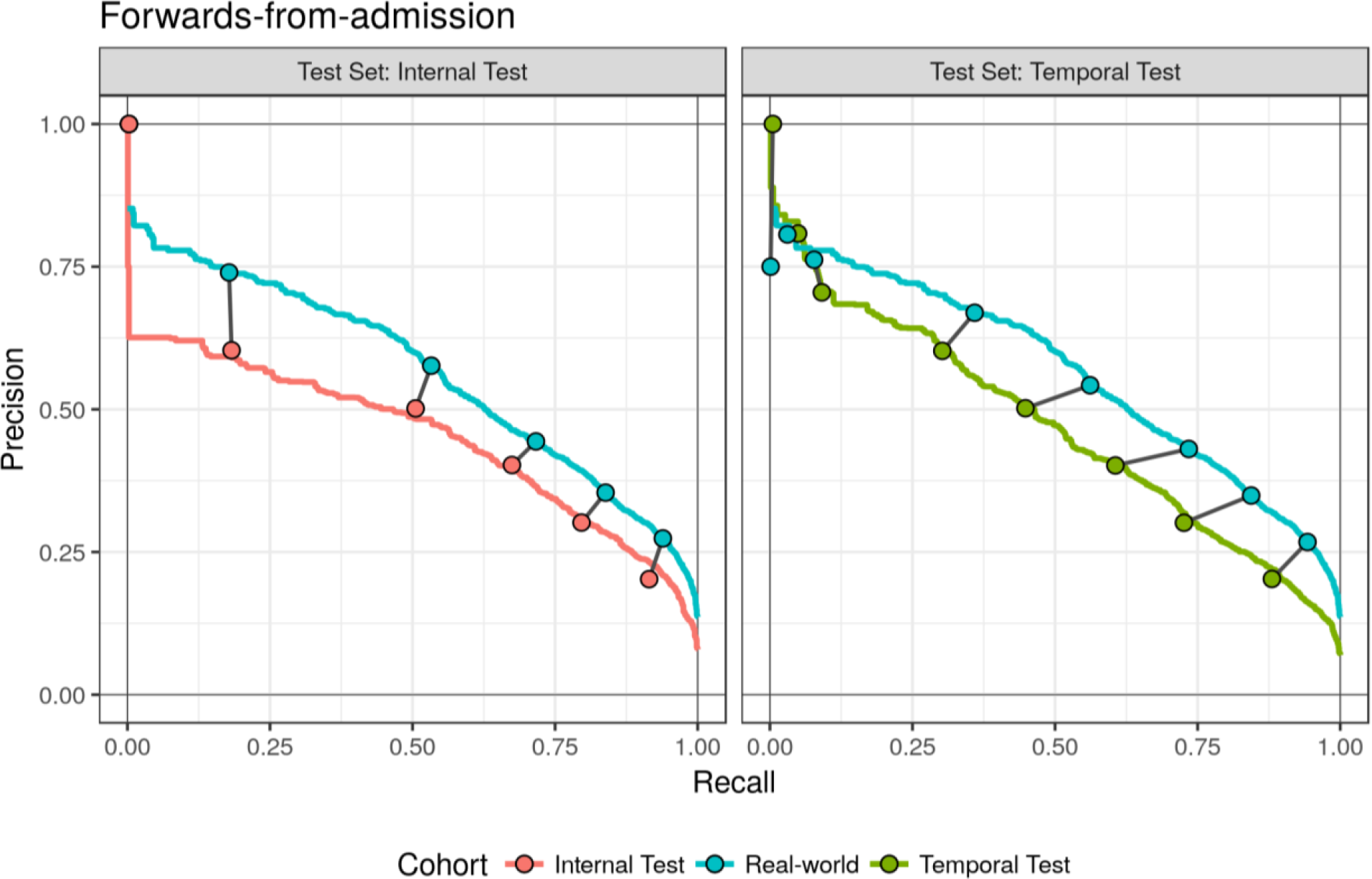
Precision and recall curves with highlighted operating points describing estimated real-world performance by the **forwards-from-admission** experimental design in comparison to the internal and temporal test sets.

### Real-world Performance

The backwards-from-outcome design excludes many patients without certain labels which improves the model’s ability to distinguish very high-risk patients from at-risk patients (for example, recall < 0.20). But, the result is a drastically smaller training cohort than the forwards-from-admission alternative. The additional sample size of the forwards-from-admission experiment **improves real-world performance by 5% AUROC and 14% AUPRC**, described in Table 2, that is **consistently and conservatively estimated with either test set**.

## DISCUSSION

### Experimental Design Considerations

Backwards-from-outcome selection limits one individual to one outcome group even if no appropriate prediction instant can be identified. In reality, patients that die are likely to be hospitalized more than once (55% of patients in our dataset), and in many cases will have long clinical histories with increasing risk. Since the forwards-from-admission design determines an outcome per admission, readmissions are accommodated and the model is penalized for premature or delayed risk estimation along each patient’s trajectory.

However, the forwards-from-admission design may introduce bias upon the temporal test set. To mitigate data leakage, all temporal test patients must have not been recently admitted—otherwise they would have been recruited for training—and therefore may be less acutely ill. A brief drop in admissions is noticeable in the death group of Figure 2 (bottom-right). A more rigorous experiment may separate training and temporal testing by a buffer time period. Moreover, the inclusion of readmissions violates the assumption that instances are independent and identically distributed.

### Simulated Deployment

The thresholding and simulated deployment presented here is overly simplified. In practice, thresholding would challenge the quality of some ‘gold-standard’ outcome labels, especially those patients predicted at relatively high-risk. Since death data is prone to missingness [17], a process including chart review or follow-up may reconcile some cases. Any improvements may be particularly helpful in the forwards-from-admission internal test set as thresholding is currently hindered by poor performance at very low recall.

In this experiment, the real-world cohort begins immediately after the temporal test set. In reality, at least 1 year will pass between development data and implementation, to collect outcomes and develop the model. Implementation will almost certainly be more difficult than this simulated deployment such that measuring generalization with a sampled test set becomes inadequate. Instead, a temporally separated test set will likely yield more realistic estimates of performance as it assesses the model’s adaptation to the future—historic transportability.

### Limitations

The results presented here consider one dataset. However, it is not unreasonable to expect this pattern of behavior in other datasets. Although we cannot predict future changes, we can be sure they will arrive. Expecting the challenges of deploying in a temporally evolving domain highlights the importance of pragmatic solutions for calibration drift [18] that may require retraining or planned re-calibration [19], but future research is required.

Comparison to previous work is unfortunately limited as only 5 years of data was available in a setting with, potentially, less continuity of care and follow-up than Avati et al. [2] (given the geographic differences between New York City and Santa Clara County). In addition, predictions were restricted to inpatient admissions for practicality, a random forest model was employed for portability and efficiency, and hospice discharges were included as a proxy for death upon observing that hospice patients were often lost to follow-up with no confirmed death date [unpublished].

Using a binary ‘gold-standard’ outcome label is common [20], however, implementation evaluation is plagued with uncertainty. Cohort selection and strict inclusion/exclusion criteria inadvertently separate the two classes, but patients identified in deployment will be lost to follow-up. Testing on a dataset that doesn’t represent the expected deployment distribution is likely optimistic.

The extreme reduction in cohort size when selecting backwards-from-outcome results in relatively small cohorts in the internal and temporal test sets, especially the death groups of 192 and 348 admissions respectively. These groups are likely inadequate for reliable thresholding within the precision recall space, especially under subsampling conditions. More testing data may improve the thresholds. However, the sanitization of data by the backwards-from-outcome design will limit the utility of any test set threshold upon deployment.

When applying machine learning to healthcare data, experimental choices are crucial, not only for their effect on validation performance but also on the real-world performance at a specific operating criterion selected to drive downstream actions. Without transparent reporting of these design choices, there is no way to discriminate between potentially disruptive new applications and those reliant on biased, idealistic results. Since some experimental designs sanitize the underlying data beyond resemblance to real-world data, these designs should be discouraged by the community; their continual use only perpetuates unrealistic expectations of machine learning and artificial intelligence that may fuel disillusionment.

### Future work

We consider a simplified scenario where we plan to evaluate the learned model for use within our institution. Before we comment on its utility elsewhere, spectrum bias [21] and other pillars of transportability [1] must be addressed. Our first step is generalizing to other hospitals within our system which contain different patient distributions with varying data collection processes that may impede generalization.

An estimate of 1 year mortality risk can only be so impactful in clinical practice. Shorter time periods may prove more useful for decision-making and to convey to the patient and their family. We plan to develop a similar model to predict shorter term mortality.

Prediction that relies on data from the day of admission is expected to encounter more implementation challenges than using entirely historical data. In particular, some data fields included here may be ‘leaking’ into the model due to data fidelity limitations within the electronic medical record. We plan to operationalize a similar model for implementation with the goal of developing clinical decision support that initiates a clinical workflow for high-risk patients.

## CONCLUSION

Evaluation of a predictive model in terms of its utility and feasibility for deployment presumes the reported performance is achievable in the real-world. In this work, we evaluated two common retrospective experimental designs employed for model development, and selected a range of thresholds across two test sets in order to compare their differences in terms of estimated model performance. Cohort selection in a forwards-from-admission design results in higher real-world performance. Moreover, a temporally separated test set yields more consistent estimates of real-world performance especially at higher precision. In all cases, migration from test set to real-world performance was observed and should be expected when deploying predictive models into clinical practice.

## Data Availability

No raw data is available. Aggregated data used to make the included tables and figures may be available upon reasonable request.

## ACKNOWLEDGEMENTS

We thank Simon Jones for early discussion and feedback.

## STUDY APPROVAL

The NYU School of Medicine IRB has determined that this work is not human subjects research and therefore IRB review was not required and not obtained.

